# Longitudinal Associations between the Built Environment, Biological Aging and Cognition

**DOI:** 10.64898/2026.10.08.26365061

**Authors:** Faye Sanders, Vilte Baltramonaityte, Marlene Staginnus, Rosa H. Mulder, Ryan L. Muetzel, Martine Vrijheid, Erin C. Dunn, Helen L. Fisher, Charlotte A.M. Cecil, Esther Walton

## Abstract

Features of urbanicity have been linked to poor psychiatric health in adulthood, but little is known about their relationship with biological age and cognition earlier in life.

Using the Avon Longitudinal Study of Parents and Children (ALSPAC), we tested associations between the built environment and epigenetic ages (EpiAge_Altum_, EpiAge_Horvath_, EpiAge_PACE_) (n=2,452), brain age (n=751), and cognition (n=12,199). Analyses were replicated in Generation R and triangulated in the Millennium Cohort Study.

The built environment was not associated with epigenetic ages (β_Altum_ range = 0.013-0.015, *p* range = .466-.689; β_Horvath_ range = -0.011-0.023, *p* range = .182-.728; β_PACE_ range = -0.045-0.013, *p* range = .131-.524) or brain age (β range = 0.022-0.028, *p* range = .337-.429). A negative association with cognition was observed in Generation R.

We provide no consistent evidence that childhood built environment exposure relates to biological age and cognition in adolescence, suggesting these relationships may emerge in later life.

---

Almost half of the global population now live in cities, defined as having at least 1,500 people per square kilometer, and a total population of at least 50,000. As a result it is becoming increasingly important to understand how urban environments shape health and child development ^1^. While a growing body of research links urbanicity to population health outcomes ^2,3^, most of this evidence is limited to adult populations. However, understanding relationships between exposure to the built environment (a component of urbanicity) during childhood and health outcomes during adolescence is particularly important, as this is a period marked by rapid biological, cognitive, and neurological change ^4^, and offers a key window for preventive intervention before major physical and mental health issues emerge.

Recent advances in biomarker research have introduced measures of biological age, which may capture variation in health, cognitive function, and mortality risk that is not explained by chronological age alone. Biological age can be derived from a wide range of biological data, but is often indexed either through DNA methylation measures, known as epigenetic age ^5,6^, or through measures obtained from neuroimaging scans, known as brain age ^7^. Detecting discrepancies between biological and chronological age in early life is particularly important as it is a strong predictor of morbidity, mortality ^8,9^, and impaired cognitive performance ^10^, and hence could guide targeting of preventative interventions.

Recent studies have linked aspects of the built environment and neighbourhood factors to epigenetic age deviations. For example, proximity to green space has been associated with multiple measures of epigenetic age *deceleration* in adults ^11^. On the other hand, poorer neighbourhood quality, which included measurements of traffic volumes, crowded sidewalks and limited green space, was also associated with multiple measures of *accelerated* epigenetic age in adults ^12,13^ and differential DNA methylation in adolescents ^14^. While these findings highlight links between some urban features and epigenetic age and DNA methylation deviations, the built environment includes a broader range of factors beyond green space and neighbourhood deprivation. For example, population density has been associated with depression, a condition closely tied to advanced biological age ^2,15^. Moreover, recent work suggests that using a general factor of the built environment may reveal stronger associations with mental health outcomes than examining isolated features ^16^. This suggests that research on biological age should consider more integrative, composite indicators of the built environment, in order to better capture relationships between the built environment and mortality and health risk.

Notably, most associations between the built environment, neighbourhood factors and epigenetic age have been tested on adults. Only two studies to date have focussed on children and adolescents. Marini and colleagues ^17^ found neighbourhood disadvantage to be associated with advanced epigenetic age in one out of two tested epigenetic clocks in children aged seven. Raffington and colleagues ^18^ detected associations between neighbourhood deprivation and the pace of epigenetic aging in children and adolescents aged between eight to 18 years old. However, it remains unclear how diverse built environment exposures collectively relate to epigenetic aging in adolescence.

Built environment research on brain age remains sparse, but there is consistent evidence identifying exposure to the built environment during childhood to be associated with brain structure cross-sectionally and longitudinally, including gray and white matter reductions ^19,20^ and decreased cortical thickness ^21^. Whilst brain structure is a component of brain age, studying brain age directly may be particularly valuable as it generally outperforms individual structural brain measures in predicting brain health and is a useful biomarker of multiple age-related diseases ^7,22^, offering greater insight into relationships between built environment exposure and overall health.

Associations between the built environment and cognitive functioning are numerous, but mixed. For example, the built environment has been linked to cognitive outcomes, with studies in adults showing urban residents outperforming rural residents in cognition, navigation, working, episodic and visuospatial memory ^23–25^. Studies in children living in urban areas, on the other hand, reported reduced cognitive functioning in comparison to their rural counterparts ^26^. These mixed findings highlight the need for further research to investigate how the built environment, particularly within urban areas, relates to cognition in younger people.

To address the above challenges, we aimed to investigate associations between a composite factor of the built environment with measures of epigenetic age, brain age and cognitive functioning in young people. We preregistered our study prior to analysis on the Open Science Framework (https://osf.io/8vdw4/files/r4yha). We hypothesised that a composite built environment factor (at age 12) would be associated 1) with both accelerated brain age and epigenetic age (in late adolescence to early adulthood) and 2) with impaired cognition (in early and late adolescence to early adulthood).

## Results

### Descriptive Statistics

In ALSPAC, the three analytic subsamples (epigenetic age, brain age, and cognition) were very consistent in terms of built environment scores (**Table 1, SM Table S1**). Age distributions differed, with the epigenetic age sample being the youngest and the cognition sample the oldest. Sex ratios also varied as expected, with the brain age sample having a higher proportion of males (71%) due to recruitment focussing on males for one of the MRI sub-studies. Across all samples, participants were mostly white with the percentage of non-white participants ranging from 2.5-4.6%. Socioeconomic status (SES) risk factors and self-reported smoking were slightly elevated in the larger cognition sample. Descriptive characteristics for the Generation R and Millennium Cohort Study (MCS) samples are also presented in Table 1. For sample overlap, see **SM Figure S1**.

**Table 1.**
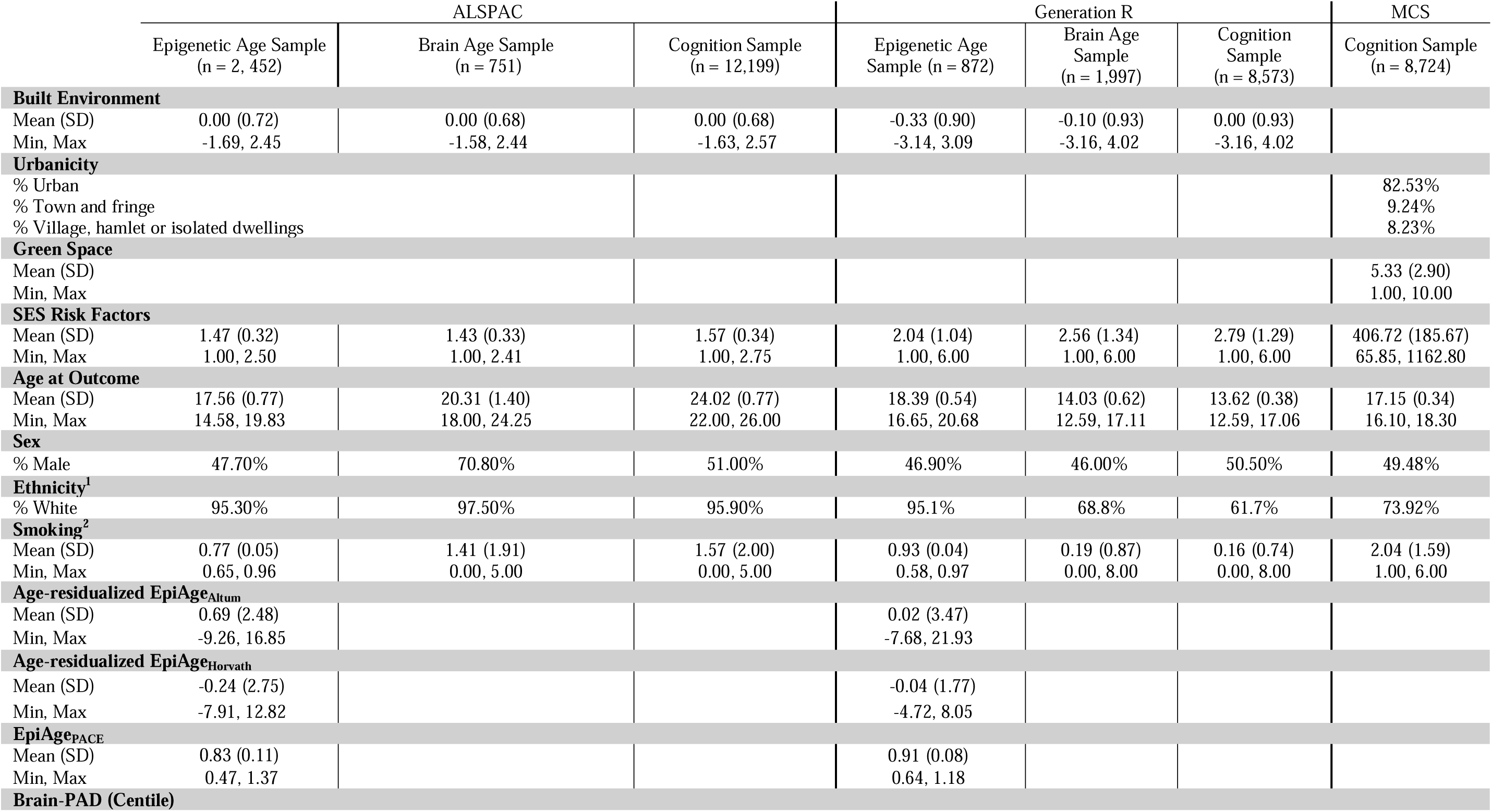
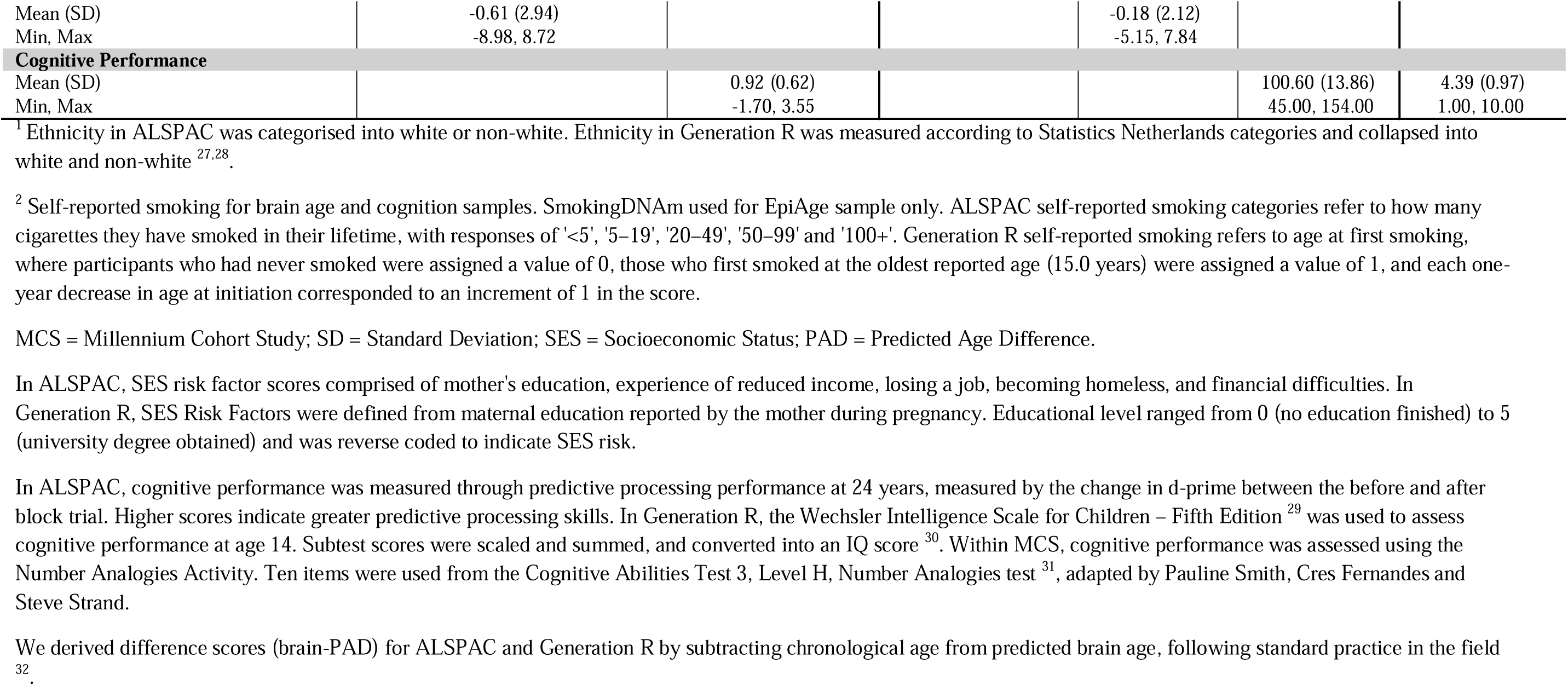
Descriptive statistics for the epigenetic age, brain age and cognition subsamples.

#### Associations between built environment factor and age-residualized EpiAge estimates

In ALSPAC, the built environment factor (measured at age 12) did not associate with either age-residualized EpiAge_Altum_ (β = 0.015, SE = 0.02, *p* = .466), age-residualized EpiAge_Horvath_ (β = 0.023, SE = 0.02, *p* = .182) or EpiAge_PACE_ (β = 0.013, SE = 0.02, *p* = .524) (see **Figure 1** and **Table 2**); these models had >95% statistical power to detect small effects ( 0.02). Although statistically insignificant, all three associations showed a consistent positive direction of effect, indicating that higher built environment factor scores may relate to slightly greater epigenetic age acceleration. In Generation R, the built environment factor (measured at age 12) did not associate with either age-residualized EpiAge_Altum_ (β = 0.013, SE = 0.03, *p* = .689), age-residualized EpiAge_Horvath_ (β = -0.011, SE = 0.03, *p* = .728) or EpiAge_PACE_ (β = -0.045, SE = 0.03, *p* = .131) and showed inconsistent directions of effect; these models had >70% power to detect small effects. A full results table including covariates (sex, cell type, batch, array [450K/EPIC], smoking_DNAm_ and socioeconomic status (SES) risk) can be seen in **SM Table S2**.

**Figure 1.**
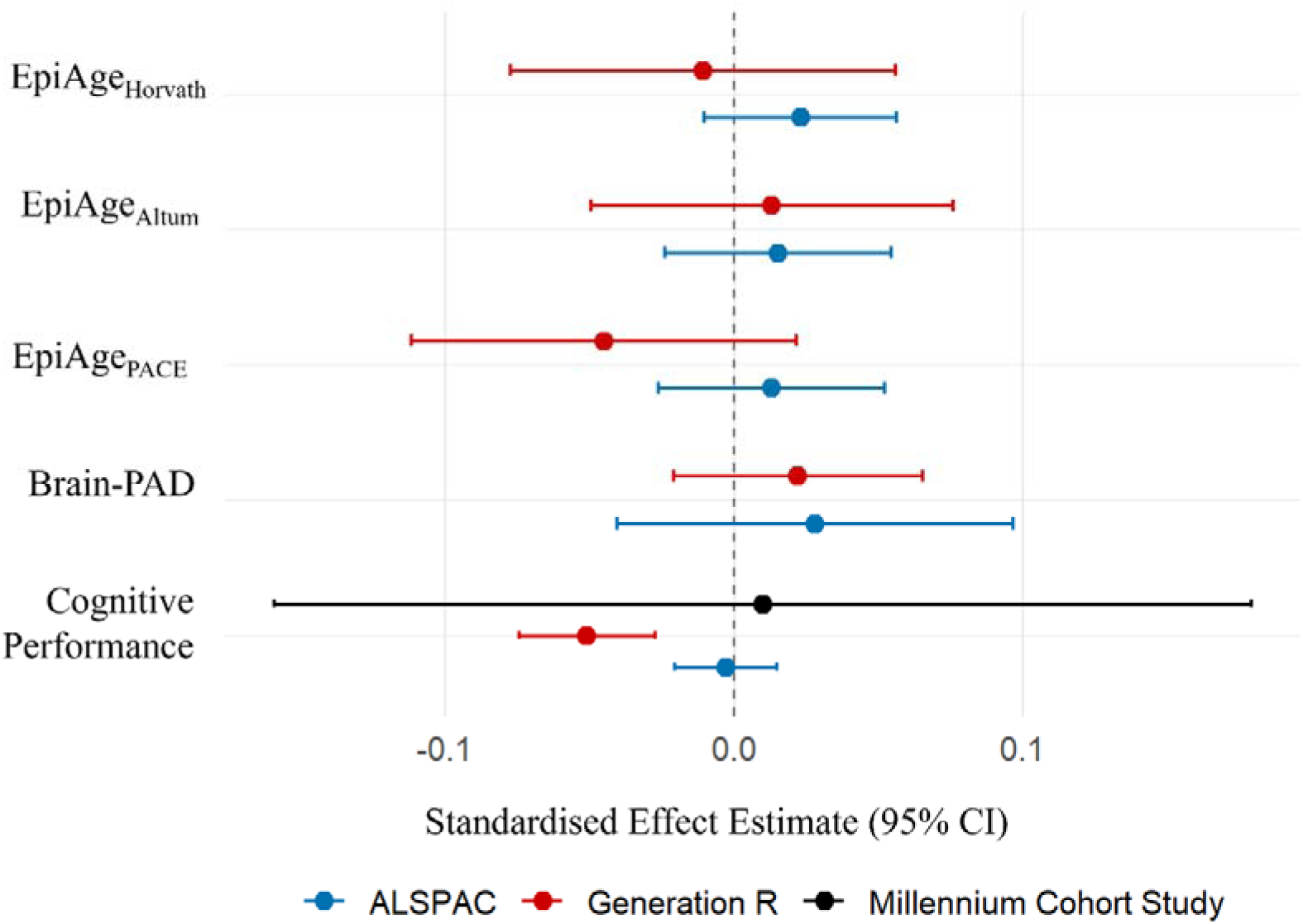
Forest plot of separate adjusted and standardised models for the associations between a built environment factor and age-residualized EpiAge, brain-PAD (predicted age difference) and cognitive performance across ALSPAC (Avon Longitudinal Study of Parents and Children), Generation R and Millennium Cohort Study. The estimate plotted for the Millennium Cohort Study is urbanicity. Epigenetic models were additionally adjusted for sex, socioeconomic status (SES) risk, smoking_DNAm_, chip/array, cell types and batch. Brain-PAD models were adjusted for sex, age, SES risk, smoking, and MRI study. Cognitive performance models were adjusted for age at assessment, sex, smoking and SES risk.

**Table 2.**
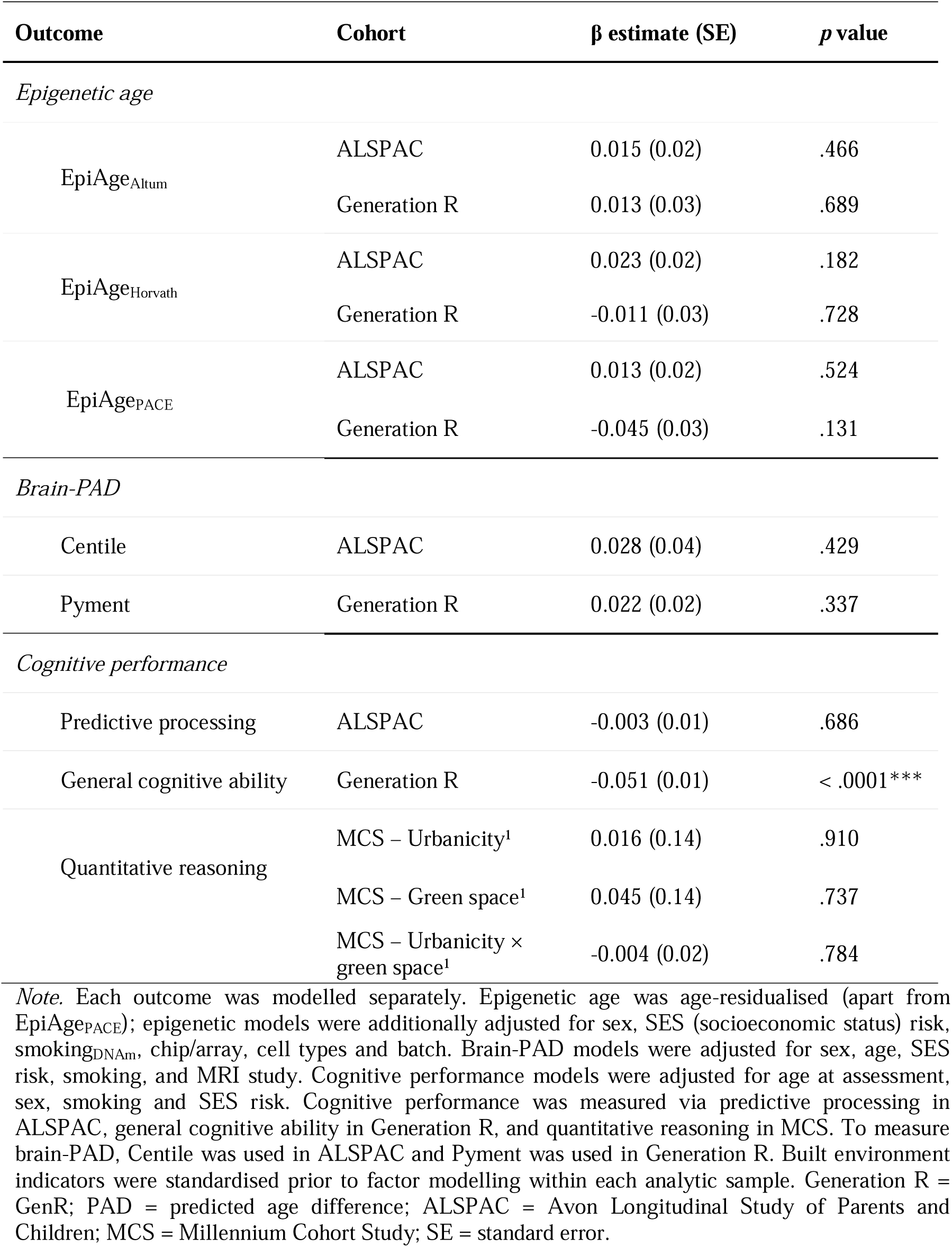
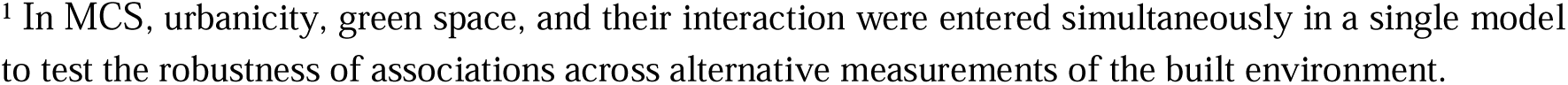
Results from separate multiple regression models testing associations between the built environment factor and epigenetic age (n_ALSPAC_ = 2,452; n_GenR_ = 872), brain-PAD (n_ALSPAC_ = 751; n_GenR_ = 1,997), and cognitive performance (n_ALSPAC_ = 12,199; n_GenR_ = 8,573; n_MCS_ = 8,724) across three cohorts.

| Outcome | Cohort | $\beta$ estimate (SE) | p value |
| --- | --- | --- | --- |
| *Epigenetic age* |  |  |  |
| EpiAge<sub>Altum</sub> | ALSPAC | 0.015 (0.02) | .466 |
|  | Generation R | 0.013 (0.03) | .689 |
| EpiAge<sub>Horvath</sub> | ALSPAC | 0.023 (0.02) | .182 |
|  | Generation R | -0.011 (0.03) | .728 |
| EpiAge<sub>PACE</sub> | ALSPAC | 0.013 (0.02) | .524 |
|  | Generation R | -0.045 (0.03) | .131 |
| *Brain-PAD* |  |  |  |
| Centile | ALSPAC | 0.028 (0.04) | .429 |
| Pyment | Generation R | 0.022 (0.02) | .337 |
| *Cognitive performance* |  |  |  |
| Predictive processing | ALSPAC | -0.003 (0.01) | .686 |
| General cognitive ability | Generation R | -0.051 (0.01) | < .0001*** |
| Quantitative reasoning | MCS – Urbanicity<sup>1</sup> | 0.016 (0.14) | .910 |
|  | MCS – Green space<sup>1</sup> | 0.045 (0.14) | .737 |
| | MCS – Urbanicity × green space<sup>1</sup> | -0.004 (0.02) | .784 |

*Note.* Each outcome was modelled separately. Epigenetic age was age-residualised (apart from EpiAge<sub>PACE</sub>); epigenetic models were additionally adjusted for sex, SES (socioeconomic status) risk, smoking<sub>DNA</sub>, chip/array, cell types and batch. Brain-PAD models were adjusted for sex, age, SES risk, smoking, and MRI study. Cognitive performance models were adjusted for age at assessment, sex, smoking and SES risk. Cognitive performance was measured via predictive processing in ALSPAC, general cognitive ability in Generation R, and quantitative reasoning in MCS. To measure brain-PAD, Centile was used in ALSPAC and Pyment was used in Generation R. Built environment indicators were standardised prior to factor modelling within each analytic sample. Generation R = GenR; PAD = predicted age difference; ALSPAC = Avon Longitudinal Study of Parents and Children; MCS = Millennium Cohort Study; SE = standard error.
<sup>1</sup> In MCS, urbanicity, green space, and their interaction were entered simultaneously in a single model to test the robustness of associations across alternative measurements of the built environment.

**Figure 2.**
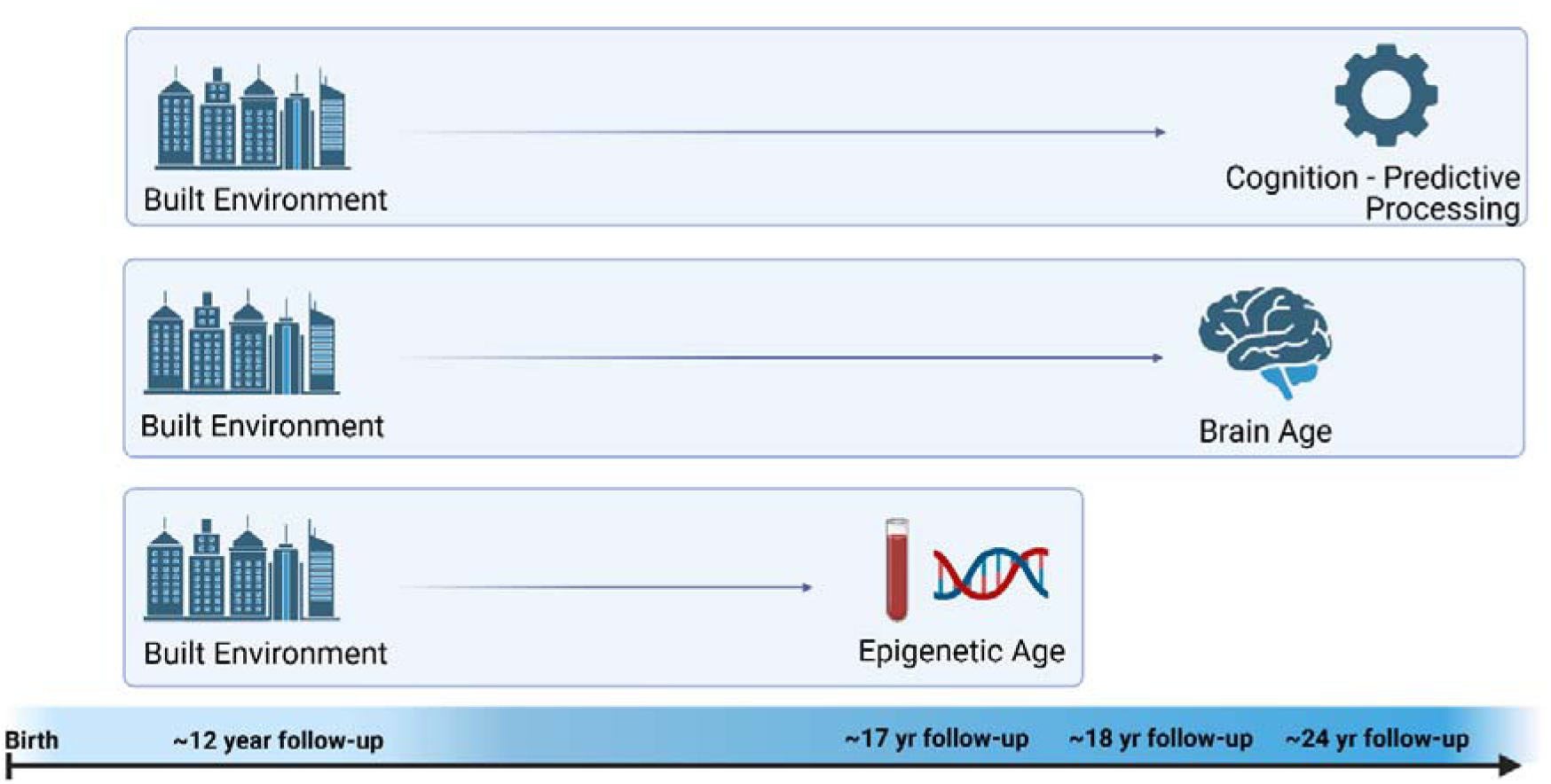
ALSPAC analysis overview. The timeline along the x axis indicates timepoints variables were measured at in ALSPAC.

#### Associations between built environment factor and brain-PAD

In ALSPAC, the built environment factor (measured at age 12) did not associate with brain-PAD (measured between ages 18-24 years) (β = 0.028, SE = 0.04, *p* = .429), covarying for sex, SES risk, smoking, age at scan and MRI study; this model had **>**70% power to detect small effects. In Generation R, the built environment factor (at age 12) did not associate with brain-PAD (measured at age 14) (β = 0.022, SE = 0.02, *p* = .337); this model had >85% power to detect small effects.

#### Associations between built environment factor and cognitive performance

In ALSPAC, the built environment factor (at age 12) was not associated with cognition (β = -0.003, SE = 0.01, *p* = .686) (see **Table 2**), covarying for age at assessment, sex, smoking and SES risk. Similarly, in MCS, the urbanicity and green space interaction was not associated with cognition (β = -0.004, SE = 0.02, *p* = .784); this model had >99% power to detect small effects. Further, urbanicity was not associated with cognition (β = 0.016, SE = 0.14, *p* = .910), nor was green space (β = 0.045, SE = 0.14, *p* = .737). In Generation R, the built environment factor (at age 12) was associated with poorer cognitive performance (β = - 0.051, SE = 0.01, *p* <.0001); this model had >99% power to detect small effects.

Generally, adjusted and sensitivity models showed a similar pattern of results across all cohorts (**SM Figure S2**). Sensitivity models included all covariates apart from smoking, as smoking is associated with the built environment, brain-PAD, cognition and age-residualized epigenetic age ^33–36^.

We conducted a sensitivity analysis in ALSPAC examining individual associations between built environment indicators and age-residualized epigenetic age, brain-PAD and cognitive performance. The pattern of associations closely matched the latent factor. Across brain-PAD, epigenetic age and cognitive performance, no single indicator showed a consistent significant association with all outcomes. Only two isolated associations reached nominal significance (walkability with EpiAge_Horvath_; facility richness with cognitive performance), but these were small and inconsistent across outcomes (**SM Table S3**).

## Discussion

The main finding of this study was that no consistent associations were found between childhood exposure to the built environment and biological age and cognition in late adolescence and early adulthood across the three longitudinal studies utilised, despite most analyses being statistically well-powered.

### Built Environment and Epigenetic Age

First, we found no clear association between childhood exposure to the built environment and several measures of epigenetic age. Although most associations were positive, none reached statistical significance. Our findings contrast with previous research in adults, which found poorer neighbourhood quality associated with epigenetic age acceleration, including the Horvath and Hannum clocks, whilst increases in green space have been associated with epigenetic age deceleration ^11–13^. However, our findings partially replicate those reported by Marini and colleagues ^17^. Although the authors of that study identified an association between neighbourhood disadvantage and the Hannum epigenetic clock in children, they similarly found no association with the Horvath clock. Taken together, these findings may suggest longer-term exposure to the built environment may be necessary for measurable associations with epigenetic age acceleration to emerge. Alternatively, whereas previous studies focused on neighbourhood quality, the broader built environment factor examined in this study may capture a more heterogeneous set of environmental characteristics, potentially obscuring associations with epigenetic ageing.

### Built Environment and Brain-PAD

Similarly, we found no clear association between childhood exposure to the built environment and brain age, contrasting with previous research detecting associations between exposure to urban environments and structural MRI features including reductions in grey matter volume ^19,20^ and cortical thickness ^21^. Although brain age estimates are derived from structural MRI measures, including grey matter volume and cortical thickness, the global nature of brain age metrics, despite predicting overall mortality better, may obscure more region-specific associations ^37^. Region-specific associations may be particularly relevant during late adolescence, when there may not yet have been sufficient time for localised structural differences to manifest as detectable differences in overall brain age. Collectively, these findings suggest that studies examining associations between the built environment and brain age during childhood and adolescence may benefit from focusing on more localised measures of brain structure.

### Built Environment and Cognitive Performance

Lastly, we found no consistent association between the built environment and cognitive performance. The null association observed in ALSPAC was not replicated in Generation R where greater exposure to built environments was associated with poorer cognitive performance. However, triangulation in MCS, using different operationalisations of both the built environment and cognitive performance, similarly found no clear association, providing some support for the null finding observed in ALSPAC ^38^. The absence of a consistent association contrasts with previous adult studies reporting urban–rural differences across multiple cognitive domains, including working memory ^23,24^. However, the negative association observed in Generation R aligns with prior findings in children linking greater urban exposure to reduced cognitive performance ^26^. Differences in how cognition was measured may explain these findings. Although the built environment variable was harmonised between ALSPAC and Generation R, the cognitive assessments differed substantially. Specifically, the cognitive measures used in ALSPAC (predictive processing) and MCS (quantitative reasoning) may have captured narrower or different domains of cognition than those most sensitive to built environment exposure, whereas the broader, general cognitive ability measure used in Generation R may have more closely reflected the relevant cognitive processes. Furthermore, cognitive performance in Generation R included a language component, meaning children who were not native Dutch speakers, and perhaps who are more likely to experience higher levels of the built environment exposure represented in our factor, may have been disadvantaged on this measure. Moreover, cognition in Generation R was measured at age 14 whereas cognition in ALSPAC was measured at age 24. The shorter interval between built environment exposure and cognitive assessment in Generation R may have reduced the opportunity for residential mobility and subsequent changes in environmental exposure. In contrast, the longer follow-up period in ALSPAC may have allowed participants to move to different environments or for any earlier effects of the built environment on cognition to diminish or recover over time. Additionally, geographical differences may have contributed to heterogeneity in findings. For example, Bristol may have greater levels of green space than Rotterdam ^39,40^. It is therefore possible that the more urbanised context of Rotterdam contributed to the negative association observed between the built environment and cognitive performance in Generation R only. Taken together, triangulation across cohorts suggests that any association between built environment exposure and adolescent cognitive performance is likely context dependent and does not generalise across different cognitive measures or settings.

### Limitations

It is also important to consider the following limitations of our findings. Whilst previous studies have used a mixture of urban and rural residents, most participants across all cohorts in our study were living in urban settings. As a result, variability in the built environment factor was limited and hence statistical power to observe associations was likely attenuated. In addition, the built environment was only measured at one timepoint. As a result, we were unable to capture cumulative exposure, residential mobility, or changes in neighbourhood environments across development, which may be particularly important when investigating long-term health and cognitive development processes ^41,42^. Secondly, biological age was estimated only in subsets of participants who had either undergone structural MRI or had DNA methylation samples processed. If factors influencing selection into these subsamples were also associated with the built environment factor, this could have biased observed associations. Lastly, apart from one third generation clock trained to measure the pace of biological ageing, most biological age measures used in the present study were trained to predict chronological age. Associations may have differed had we used second-generation clocks ^43^, which are designed to predict health outcomes and may therefore be more sensitive to environmental exposures. However, the application of these measures in younger cohorts remains limited, as they have largely been developed and validated in adult populations.

### Conclusions

To conclude, this study aimed to investigate relationships between childhood exposure to the built environment and biological age and cognition in late adolescence and early adulthood. Results indicated no clear association between the built environment with multiple measures of biological age and cognitive performance. These findings suggest that associations between the built environment and biological ageing may emerge later in adulthood. They further indicate that associations between the built environment and young people’s cognitive outcomes may be context-dependent and sensitive to differences in cohort or location characteristics and outcome measurement. Further research using more regionally-specific measures of brain structure and standardised assessments of general cognitive ability across diverse settings may be needed to improve understanding of these relationships.

## Methods

### Participants

#### Primary Sample

Participants were children from the Avon Longitudinal Study of Parents and Children (ALSPAC) cohort ^44,45^. Pregnant women resident in Avon, UK with expected dates of delivery 1st April 1991 to 31st December 1992 were invited to take part in the study and the initial number of pregnancies enrolled was 14,541. Of the initial pregnancies, there was a total of 14,676 foetuses, resulting in 14,062 live births and 13,988 children who were alive at 1 year of age.

We included children who had available at least 50% built environment data at study enrolment and at least one of the following outcomes measured between ages 8 and 24: DNA methylation, structural MRI, or cognitive performance. One outlier was removed in the epigenetic age sample as two out of three epigenetic age estimates were considerably higher than the rest of the sample (15.03 standard deviations above the mean for age-residualized EpiAge_Altum_ and 9.46 standard deviations above the mean for age-residualized EpiAge_Horvath_).

Missing data for the exposure, covariates, and non-biological outcomes (i.e., cognitive performance) were imputed (see **SM section 1.5**). To avoid potential biases arising from genetics and shared family environments, only the first-born twin in the case of twin-pairs and first study child in the case of siblings are included in our sample.

Ethical approval for the study was obtained from the ALSPAC Ethics and Law Committee and the Local Research Ethics Committees and Department of Psychology at the University of Bath. The study website (https://www.bristol.ac.uk/alspac/researchers/our-data/) contains details of all data available through a fully searchable data dictionary and variable search tool.

#### Replication and Triangulation Samples

The Generation R Study is a population-based cohort on health and development from fetal life onwards ^28,46^. Pregnant women residing in the study area of Rotterdam, the Netherlands, with an expected delivery date between April 2002 and January 2006 were invited to participate. The study has been approved by the Medical Ethical Committee of Erasmus MC, University Medical Center Rotterdam. Written informed consent was obtained from parents of all children ^28,46^. In total, 9,778 mothers were enrolled in the study and gave birth to 9,749 live-born children. Of these, 9,258 children had at least 50% of the built environment variables available at 12 years of age. After randomly selecting one sibling from each pair or set of siblings, 8,573 children remained in the sample of the cognitive performance analysis. Of these, 1,997 children also had MRI data available at 14 years and were included in the BrainAge analysis. Second, of the 8,573 children included in the cognitive performance analysis, 872 children also had DNA methylation data available at 18 years of age and were included in the EpiAge analysis.

The Millennium Cohort Study (MCS) is a prospective study of the social, economic and health-related circumstances of children born in the UK between September 2000 and January 2002 ^47,48^. The original cohort comprised 18,818 children (72% of those approached). Ethical approval was obtained for all waves of the Millennium Cohort Study through the National Health Service (NHS) Research Ethics Committee (REC).

To the best of our ability, we harmonized exclusion criteria and imputation protocol across samples.

## Measures

### Exposure

#### Built Environment

In ALSPAC and Generation R, we used a latent built environment factor at age 12, already associated with depressive symptoms in adulthood ^16^, derived from multiple environmental indicators including building density, population density, connectivity density, green space, facility richness and walkability, as generated in Sanders et al. ^16^. Other exposures associated with built environments, including air pollution and traffic, were not included or adjusted for, as these exposures or potential mediators may be downstream consequences of density related features. Adjusting for them would risk removing meaningful variance that is part of the built environment pathway itself that we intended to capture ^49^. Greater scores on the built environment factor indicate more urbanised areas (e.g., greater population density, less green space). Measures were harmonized through the Horizon 2020 LifeCycle project ^50,51^ and geo-coded to 1 m resolution ^52^. In MCS, green space and urbanicity were available for use at age 11, enabling us to investigate triangulation. Urbanicity in MCS was defined according to the 2004 Office for National Statistics Urban/Rural definitions, measured through characteristics including population density and settlement morphology ^53^. See **SM section 1.2** for full methodology and **SM Table S4** for a comparison of the factor loadings between ALSPAC and Generation R.

Covariates included self-reported smoking (age 17 ALSPAC; age 14 Generation R), smoking_DNAm_ (age 17 ALSPAC; age 18 Generation R) (DNAm_cg05575921_), cell type proportions, sex, SES risk factors, batch effects, age at outcome and MRI study. A detailed explanation of covariates can be seen in **SM section 1.3**.

### Outcomes

#### Epigenetic Age Acceleration

In ALSPAC and Generation R, we derived three measures of epigenetic age including:

1) EpiAge_Altum_ ^54^ (https://github.com/rsinghlab/AltumAge)
2) EpiAge_Horvath_ ^55^, a skin and blood clock trained on fibroblasts, blood, skin, buccal, saliva, and related tissues (https://github.com/yiluyucheng/dnaMethyAge)
3) EpiAge_PACE_ which is a rate-rather than state-based measure that predicts the pace of aging ^56^. (https://github.com/danbelsky/DunedinPACE)

DNA methylation data from blood samples was used to derive these measures. See **SM section 1.4** for details on DNA methylation preprocessing steps. These measures were selected as they have been trained on young people (EpiAge_Altum_: n = 8050, -0.8-112.0yrs; EpiAge_Horvath_: n = 896, 0-94yrs; EpiAge_PACE_: n = 817, 26-45yrs), covering first and third generation of clocks. EpiAge_Horvath_ performs well when predicting age in ALSPAC adolescents and other population-based cohorts ^54,57,58^.

In line with the field, we derived age-residualized epigenetic age scores by regressing epigenetic age on chronological age and retaining the residuals for state-based measures ^58^. Positive residuals indicate an epigenetic age older than expected for chronological age, whereas negative residuals indicate a younger-than-expected epigenetic age.

#### Brain-PAD

In ALSPAC, structural MRIs were obtained on a subset of participants (n=958, 18 to 24 years) in three ALSPAC neuroimaging studies ^59^. Using the MRI data, a measure of brain age difference was derived in early adulthood. BrainAge_Centile_ uses multivariate fractional polynomial regression applied to morphometric measures including intracranial volume, mean cortical thickness, total surface area, thickness and surface area from 34 bilateral cortical regions (Desikan–Killiany atlas), and volumes of 14 subcortical structures (ASEG atlas) ^60^, trained on individuals aged between 5-40 years. BrainAge_Centile_ is selected as it performs well in predicting chronological age in ALSPAC adolescents ^57,58,61^.

In Generation R, magnetic resonance imaging (MRI) data were collected at 14 years of age ^62^. For further details see **SM section 1.4**. Using the MRI data, a measure of brain-PAD was derived in adolescence from chronological age at scan. BrainAge_Pyment_ was selected in Generation R over BrainAge_Centile_ due to data governance restrictions. BrainAge_Pyment_ uses a convolutional neural network model to estimate brain age from T1-weighted structural MRI. BrainAge_Pyment_ is trained on a large multisite dataset of raw structural MRI scans including males (n = 25,827) and females (n = 27,715) aged between 3-95 years old ^63^.

We derived difference scores (brain-PAD) for ALSPAC and Generation R by subtracting chronological age from predicted brain age, following standard practice in the field ^32^. Positive brain-PAD values indicate that an individual’s brain appears older than expected for their chronological age, whereas negative values indicate that an individual’s brain appears younger than expected.

#### Cognitive Performance

The use of prior knowledge in perceptual decision-making is considered an important aspect of the cognitive processes underlying working memory and fluid intelligence ^64,65^. In ALSPAC, predictive processing performance was assessed at 24 years, measured by the change in *d*-prime between the before and after blocks. The task involved first viewing ambiguous black-and-white images and being asked whether they could see a person or animal. Participants were then shown colour versions of the images and repeated the task to see whether this new information changed what they could identify in the original images. *D*-prime represents the influence of knowledge on perception using signal detection theory ^66,67^. Higher scores indicate greater predictive processing skills.

In Generation R, the Wechsler Intelligence Scale for Children – Fifth Edition (WISC-V) ^29^ was used to assess cognitive performance during the research visit at age 14. The scale included Matrix reasoning to measure fluid reasoning, Digit Span for working memory, Coding for processing speed, and Vocabulary for verbal comprehension. Subtest scores were scaled and summed, and converted into an IQ-score ^30^.

Within MCS, cognitive performance was assessed using the Number Analogies Activity at age 17. Ten items were used from the Cognitive Abilities Test 3, Level H, Number Analogies test ^31^, adapted by Pauline Smith, Cres Fernandes and Steve Strand. Quantitative reasoning forms a part of many modern intelligence batteries, including the Wechsler scales ^29^.

## Statistical Analysis

We first tested associations in ALSPAC between the built environment factor at age 12 and (1) age-residualized epigenetic ages at 17 (adjusting for sex, cell type, batch, array [450K/EPIC], smoking_DNAm_ and SES risk), (2) brain-PAD at 18–24 years (sex, SES risk, smoking, age at scan, MRI study), and (3) predictive processing at 24 years (age at assessment, sex, smoking, SES risk). Built environment indicators were standardised prior to factor modelling within each analytic sample. Because smoking is associated with the built environment, Brain-PAD, cognitive performance and age-residualized epigenetic age ^33–36^, and may act as a mediator, we conducted sensitivity analyses excluding smoking/smoking_DNAm_ from our models.

We then sought replication and triangulation in Generation R and MCS. Built environment variables were harmonised across ALSPAC and Generation R via the LifeCycle Project-EU Child Cohort Network ^50^, whereas green space and urbanicity were available in MCS, allowing us to examine whether any replicated associations were robust across alternative operationalisations of the built environment.

In Generation R, we tested associations between the built environment factor at age 12, generated and standardised across the full cohort, and (1) age-residualized epigenetic age at 18 (sex, cell type, batch, smoking_DNAm_, SES risk), (2) brain-PAD at 14 years (sex, SES risk, smoking, age at scan), and (3) cognitive performance at 14 years (age at assessment, sex, smoking, SES risk). In MCS, we did not construct the same built environment factor. Instead, as we had two measures including urbanicity and green space, enabling us to examine triangulation. To do so, urbanicity, green space, and their interaction were entered simultaneously in a single model, adjusting for age, sex, SES risk, and smoking. This provided an alternative measurement to the multidimensional built environment factor examined in the other cohorts.

## Acknowledgements

We are extremely grateful to all the families who took part in this study, the midwives for their help in recruiting them, and the whole ALSPAC team, which includes interviewers, computer and laboratory technicians, clerical workers, research scientists, volunteers, managers, receptionists and nurses.

The Generation R Study is conducted by Erasmus MC, University Medical Center Rotterdam in close collaboration with the School of Law and Faculty of Social Sciences of Erasmus University Rotterdam, the Municipal Health Service Rotterdam area, Rotterdam, the Rotterdam Homecare Foundation, Rotterdam and the Stichting Trombosedienst & Artsenlaboratorium Rijnmond (STAR-MDC), Rotterdam. We gratefully acknowledge the contribution of children and parents, general practitioners, hospitals, midwives and pharmacies in Rotterdam. The generation and management of the Illumina 450K methylation array data (EWAS data) for the Generation R Study was executed by the Human Genotyping Facility of the Genetic Laboratory of the Department of Internal Medicine, Erasmus MC, the Netherlands. We thank Mr. Michael Verbiest, Ms. Mila Jhamai, Ms. Sarah Higgins, Mr. Marijn Verkerk and Dr. Lisette Stolk for their help in creating the EWAS database. We thank Dr. A. Teumer for his work on the quality control and normalization scripts. The CLS cohorts, including the Millennium Cohort Study, are only possible due to the commitment and enthusiasm of their participants, their time and contribution is gratefully acknowledged.

## Data Availability

The study website (https://www.bristol.ac.uk/alspac/researchers/our-data/) contains details of all data available through a fully searchable data dictionary and variable search tool. Generation R data are available upon reasonable request to the director of the Generation R Study, subject to local, national and European rules and regulations. Millennium Cohort Study data are available from the UK Data Service to registered users.

## Code Availability

The code for this analysis can be found at https://github.com/fsanders1/built-environment-biological-age.

## Funding

The UK Medical Research Council and Wellcome (Grant ref: 217065/Z/19/Z) and the University of Bristol provide core support for ALSPAC. This publication is the work of the authors and FS and EW will serve as guarantors for the contents of this paper. A comprehensive list of grants funding is available on the ALSPAC website ^68^.

EW and VB are funded from UK Research and Innovation (UKRI) under the UK government’s Horizon Europe/ERC Frontier Research Guarantee [BrainHealth, grant number EP/Y015037/1]. ED and EW are also funded from the National Institute of Mental Health of the National Institutes of Health (award number R01MH113930; PI Dunn).

FS is supported by a scholarship from the EPSRC Centre for Doctoral Training in Advanced Automotive Propulsion Systems (AAPS), under the project EP/S023364/1.

C.A.M.C. is supported by the European Union’s Horizon Europe Programme (STAGE project, grant agreement no.101137146) and by the European Union’s HorizonEurope Research and Innovation Programme (FAMILY, grant agreement No 101057529; HappyMums, grant agreement No 101057390) and the European Research Council (TEMPO; grant agreement No 101039672). This research was conducted while C.A.M.C. was a Hevolution/AFAR New Investigator Awardee in Aging Biology and Geroscience Research.

RLM is supported by the European Union’s HorizonEurope Research and Innovation Programme (FAMILY, grant agreement No 101057529) and the Netherlands Organization for Health Research (Vidi).

HLF is part supported by the UK Economic and Social Research Council (ESRC) Centre for Society and Mental Health at King’s College London [ES/S012567/1; UKRI861].

The general design of The Generation R Study was made possible by financial support from Erasmus MC, Rotterdam, Erasmus University Rotterdam, the Netherlands Organization for Health Research and Development (ZonMW), the Netherlands Organization for Scientific Research (NWO), the Ministry of Health, Welfare and Sport and the Ministry of Youth and Families. The EWAS data were funded by a grant from the Netherlands Genomics Initiative (NGI)/Netherlands Organisation for Scientific Research (NWO) Netherlands Consortium for Healthy Aging (NCHA; project nr. 050-060-810), by funds from the Genetic Laboratory of the Department of Internal Medicine, Erasmus MC, by a grant from the National Institute of Child and Human Development (R01HD068437), and by a Helmholtz International Fellow Award HIFA-0174-RA-30/19 to JFF. Generation R Neuroimaging infrastructure were supported in part by funds from the Sophia Foundation awarded to R.L.M.

ISGlobal acknowledges support from the grant CEX2023-0001290-S funded by MCIN/AEI/https://doi.org/10.13039/501100011033, and support from the Generalitat de Catalunya through the CERCA Program. The study received funding from the Horizon 2020 research and innovation programme (grant agreement No. 874583 - ATHLETE project; grant agreement No. 733206 - LifeCycle project).

The Economic and Social Research Council funds the Centre for Longitudinal Studies (CLS) Resource Centre (ES/W013142/1) which provides core support for the CLS cohort studies, including the Millennium Cohort Study. While the CLS Resource Centre makes these data available, CLS does not bear any responsibility for the analysis or interpretation of these data by researchers.

## Competing interests

The authors declare no competing interests.

## Notes

### Competing Interest Statement

The authors have declared no competing interest.

### Author Declarations

The Ethics and Law Committee of The Avon Longitudinal Study of Parents And Children gave ethical approval for this work (IDs: E1808, 49/89 and 90/8). The Research Ethics Committees of University of Bristol gave ethical approval for this work. The Department of Psychology Research Ethics Committee of the University of Bath gave ethical approval for this work (6992-9812).

