## Supplementary Materials for "Longitudinal Associations between the Built Environment, Biological Aging and Cognition"

**Table of Contents**

| **Supplementary item** | **Page** |
| --- | --- |
| **SM Table S1.** Differences between the three analytic sub-samples within ALSPAC | 2 |
| **SM Figure S1.** Venn diagram displaying sample overlap in ALSPAC | 2 |
| **SM Table S2.** Multiple regression results for Epigenetic Age models | 3 |
| **SM Figure S2.** Forest plot of adjusted, unadjusted and sensitivity models | 4 |
| **SM Table S3.** Sensitivity analyses of individual built environment indicators | 5 |
| **Supplementary Section 1.2.** Built Environment Measures | 5 |
| **Supplementary Section 1.3.** Covariates | 7 |
| **Supplementary Section 1.4.** DNA methylation and MRI data processing | 10 |
| **Supplementary Section 1.5.** Missing data imputation | 11 |
| **SM Table S4.** Built Environment Factor Loadings in ALSPAC and Generation R | 13 |
| **References** | 14 |

**SM Table S1**

*Differences between the three analytic sub-samples within ALSPAC.*

| Variable | Statistic | *P* value | Effect Size | Post-Hoc comparisons |
| --- | --- | --- | --- | --- |
| Built Environment | F = 0.00 | 1.000 | - | - |
| SES Risk Factors | F = 144.50 | <0.001 | η² = 0.02 | Cognition sample differed from the Brain and Epigenetic samples; Brain and Epigenetic samples did not differ |
| Age at Outcome | F = 70235.00 | <0.001 | η² = 0.90 | All pairwise comparisons significant |
| Sex | χ² = 127.51 | <0.001 | V = 0.09 | All pairwise comparisons significant |
| Ethnicity | χ² = 11.93 | 0.0179 | V = 0.02 | All pairwise comparisons significant (very small effect) |
| *Note*. SES = Socioeconomic Status. | | | | |

**SM Figure S1**

*Venn diagram displaying sample overlap in ALSPAC.*

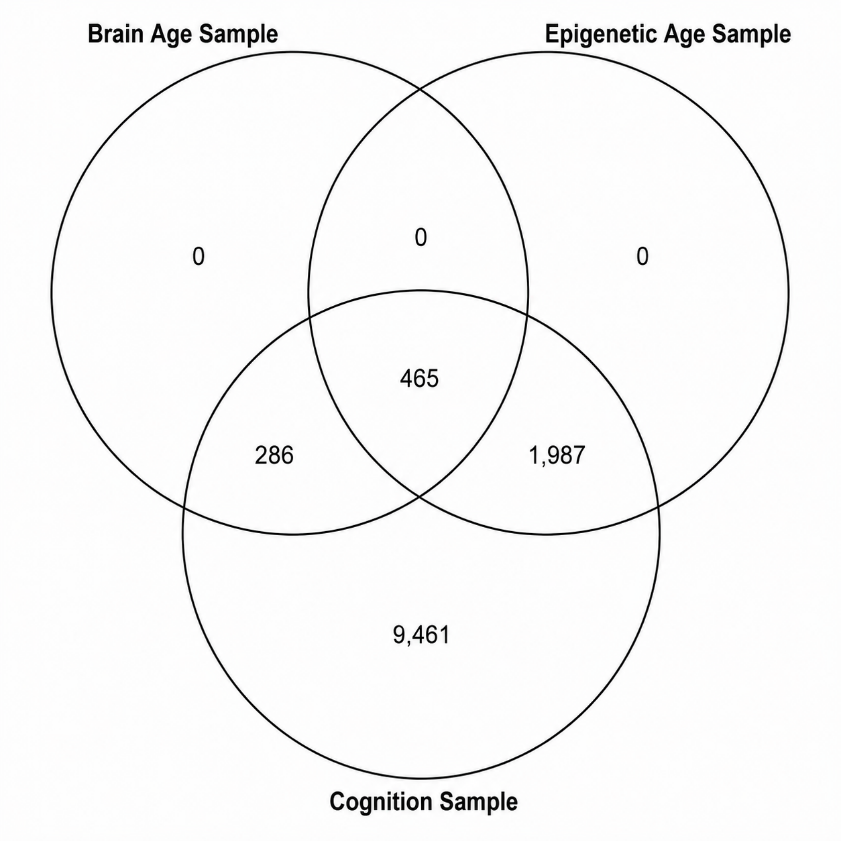

**SM Table S2**

*Multiple regression results for Epigenetic Age models (ALSPAC; n = 2,452) (Generation R; n = 872), including covariates.*

|  | **Altum EAA** | | | | **Horvath EAA** | | | | **PACE** | | | |
| --- | --- | --- | --- | --- | --- | --- | --- | --- | --- | --- | --- | --- |
|  | **ALSPAC** | | **Generation R** | | **ALSPAC** | | **Generation R** | | **ALSPAC** | | **Generation R** | |
|  | β estimate (SE) | *p* value | β estimate (SE) | *p* value | β estimate (SE) | *p* value | β estimate (SE) | *p* value | β estimate (SE) | *p* value | β estimate (SE) | *p* value |
| Built Environment Factor | 0.015  (0.02) | .466 | 0.013 (0.03) | .689 | 0.023 (0.02) | .182 | -0.011 (0.03) | .728 | 0.013 (0.02) | .524 | -0.045 (0.03) | .131 |
| Sex | 0.085 (0.104) | .417 | 0.695 (0.239) | .004** | 0.296 (0.100) | .004** | 0.653 (0.128) | < .0001*** | 0.032 (0.004) | < .0001*** | 0.013 (0.006) | .017* |
| SES Risk | -0.009 (0.02) | .672 | -0.003 (0.03) | .930 | -0.026 (0.02) | .142 | 0.024 (0.03) | .486 | 0.073 (0.02) | < .0001*** | 0.013 (0.03) | .631 |
| Smoking_DNAm_ | 0.071  (0.02) | .001** | -0.089 (0.03) | .003** | 0.037 (0.02) | .037* | -0.037 (0.03) | .246 | -0.003 (0.02) | 0.861 | -0.015 (0.03) | .622 |
| Chip | 0.444 (0.107) | < .0001*** |  |  | -0.296 (0.100) | < .0001 *** |  |  | -0.005 (0.004) | 0.27 |  |  |
| Cell type (B cell) | 0.065 (4.021) | .987 | 22.755 (6.649) | < .001*** | 31.277 (3.869) | < .0001*** | 0.807 (3.548) | .820 | 0.943 (0.159) | < .0001*** | -0.460 (0.155) | .003** |
| Cell type (CD4T) | 3.087 (5.326) | .563 | -6.686 (3.082) | .030* | 41.855 (5.124) | < .0001*** | -3.913 (1.645) | .017* | 0.408 (0.211) | 0.056 | -0.270 (0.072) | < .001*** |
| Cell type (CD8T) | 8.708 (5.603) | .123 | -4.514 (2.952) | .126 | 41.744 (5.391) | < .0001*** | -4.757 (1.575) | .003** | 0.554 (0.222) | .014* | 0.062 (0.069) | .367 |
| Cell type (Gran) | 5.808 (5.167) | .264 |  |  | 47.309 (4.972) | < .0001*** |  |  | 0.980 (0.205) | < .0001*** |  |  |
| Cell type (Mono) | 3.720 (5.649) | .512 | 3.338 (5.603) | .551 | 51.137 (5.435) | < .0001*** | 3.904 (2.990) | .192 | 1.226 (0.224) | < .0001*** | 0.165 (0.130) | .205 |
| Cell type (NK) | 11.556 (4.784) | .018* | 19.490 (2.868) | < .0001*** | 50.347 (4.603) | < .0001*** | 10.387 (1.530) | < .0001*** | 0.424 (0.189) | .027* | -0.343 (0.067) | < .0001*** |

*Note.* These analyses also adjusted for 10 surrogate variables of batch for ALSPAC, and 29 sample plates for Generation R. SES = Socioeconomic Status; EAA = Epigenetic Age Acceleration.

**SM Figure S2**

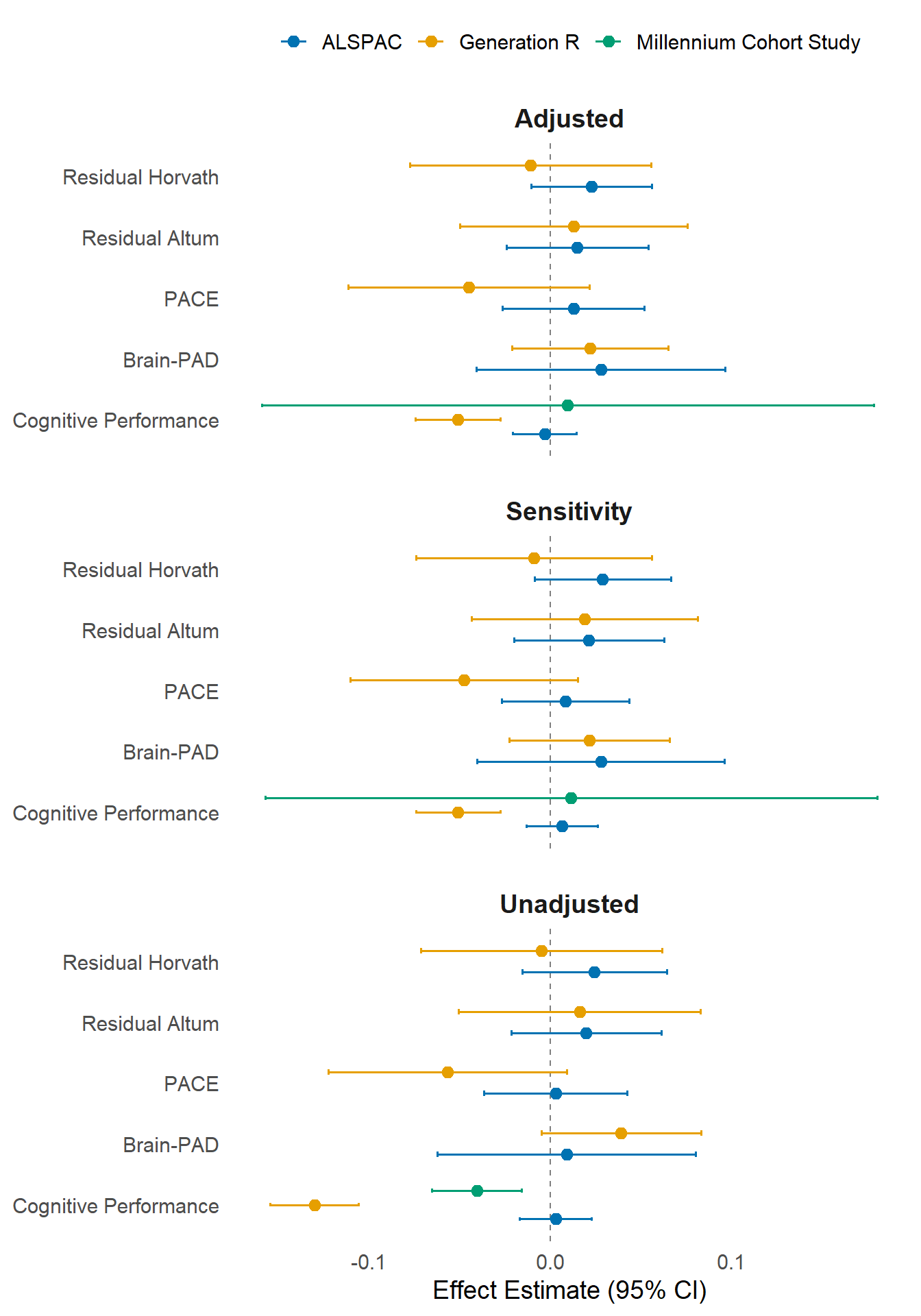
*Forest plot of adjusted, unadjusted and sensitivity models for the associations between a built environment factor and epigenetic ages, brain age and cognition across ALSPAC, Generation R and Millennium Cohort Study. Unadjusted models did not include any covariates. Sensitivity models removed smoking as a covariate from adjusted models. MCS adjusted and sensitivity models included green space and urbanicity x green space interaction term as covariates.*

**SM Table S3.**

*Sensitivity analyses in ALSPAC analysing individual associations between built environment indicators and each analytical outcome.*

|  | Brain-PAD β (SE), *p* | EpiAgeHorvath β (SE), *p* | EpiAgeAltum β (SE), *p* | EpiAgePACE β (SE), *p* | Cognitive performance β (SE), *p* |
| --- | --- | --- | --- | --- | --- |
| Building Density | 0.031 (0.04), *p* = .379 | 0.007 (0.02), *p* = .681 | 0.012 (0.02), *p* = .576 | −0.006 (0.02), *p* = .739 | 0.012 (0.01), *p* = .222 |
| Connectivity Density | 0.043 (0.04), *p* = .226 | 0.016 (0.02), *p* = .376 | 0.015 (0.02), *p* = .468 | 0.003 (0.02), *p* = .886 | 0.003 (0.01), *p* = .732 |
| Facility Richness | 0.012 (0.04), *p* = .726 | 0.031 (0.02), *p* = .085 | 0.015 (0.02), *p* = .467 | 0.011 (0.02), *p* = .544 | 0.020 (0.01), *p* = .047* |
| Green Space | −0.029 (0.04), *p* = .411 | −0.023 (0.02), *p* = .200 | −0.026 (0.02), *p* = .214 | −0.029 (0.02), *p* = .109 | 0.003 (0.01), *p* = .742 |
| Population Density | 0.013 (0.04), *p* = .705 | 0.011 (0.02), *p* = .542 | 0.010 (0.02), *p* = .639 | −0.004 (0.02), *p* = .805 | 0.005 (0.01), *p* = .615 |
| Walkability | −0.007 (0.04), *p* = .838 | 0.037 (0.02), *p* = .040* | 0.010 (0.02), *p* = .651 | 0.003 (0.02), *p* = .869 | 0.006 (0.01), *p* = .537 |

Supplementary Section 1.2

Built Environment Measures

In ALSPAC and Generation R, a built environment latent factor comprised of the following indicators. For information on how this factor was modelled, please see our related paper ^1^. In the current study, we conducted exploratory factor analyses (maximum likelihood extraction with oblimin rotation) which indicated a dominant first factor (mean eigenvalue = 3.69), accounting for 54.3% of total variance across all three ALSPAC samples. In a sensitivity analysis, this latent factor correlated with the index of multiple deprivation (measured at age 12) between 0.29 and 0.33 across all ALSPAC samples. The latent factor was first extracted, and then linear regression methods were run in R Studio.

*Building Density*

Building density was measured at age 12 by the number of m^2^ of buildings per km^2^. A 300m buffer score was used, representing the average building density within a 300m radius of the home address. A 300m buffer is commonly used in literature, however there are high correlations amongst different distance buffers and health behaviours ^2–5^. A 300m buffer strikes a balance between capturing a sufficient portion of the built environment with a likelihood of the participant interacting with that portion of their environment.

*Connectivity Density*

Connectivity density at a 300m buffer was measured at age 12. This is the number of road intersections, where two or more roads connect in some way, either through meeting or diverging ^6^, per km^2^.

*Facility Richness*

Facility richness at age 12 was measured as the number of different facility types in the 300m buffer (e.g., educational, health) available divided by the maximum potential number of facility types. It was defined through a score of 0 to 1, where greater scores indicate greater facility richness.

*Walkability*

An average walkability index score (0-1) at age 12 was measured at a 300m buffer. The score was defined as the mean of deciles for facility richness index, connectivity density, land use diversity and population density ^7,8^. The walkability index contained some components of built environment features included in the analysis, such as facility richness and population density. However, it is recommended by Descarpentrie and colleagues ^4^ to use both the walkability scores and the component variables (facility richness index etc.) in order to capture any potential information losses that can occur from transforming data into a single composite score. Greater scores of walkability are strongly associated with increased walking behaviour ^9^.

*Green Space*

Green space was quantified using the Normalized Difference Vegetation Index at age 12. The 500m buffer was selected as this was the maximum buffer radius available, and greater NDVI buffer sizes have been found to have stronger associations with health than smaller sizes ^10^. Greater buffer sizes are also important as individuals often report travelling substantially further from their home addresses specifically to reach higher quality green spaces, particularly for recreational trips ^11^. Scores range from -1 to +1, with greater scores indicating more green space.

*Population Density*

Population density, the number of inhabitants per km^2^, was measured at age 12.

*Millennium Cohort Study Built Environment Indicators*

Green space (at age 11) was measured through land-use percentile data at the lower layer super output area (LSOA) level. Land use was measured using the Generalised Land Use Database ^12^ and captures both public and private green spaces larger than 5 m^2^ in an area, excluding domestic gardens. This data was only available for participants living in England. Urbanicity (at age 11) was measured using the 2004 Urban/Rural definitions by the Office for National Statistics ^13^. On the super output area (SOA) level, areas are classed as either ‘Urban (population > 10,000)’, ‘Town and fringe’ or ‘Village, hamlet or isolated dwellings’.

Supplementary Section 1.3

Covariates

*Smoking_DNAm_*

A well-validated proxy for smoking, which is an epigenetically predicted measure, smoking_DNAm_ (based on DNAm_cg05575921_) measured at age 17 in ALSPAC ^14^ and age 18 in Generation R. Exposure to smoking is critical to account for due to its associations with epigenetic age, brain age and cognition ^15,16^. Using smoking_DNAm_ provides a more accurate measure of tobacco exposure, capturing both active and passive smoking, and helps to address underreporting challenges associated with self-reported smoking in adolescence ^14^.

*Smoking*

For BrainAge and cognitive functioning samples, self-reported smoking was used instead. In ALSPAC, this was measured at age 17. This was as most participants within these samples did not have DNA methylation data to extract smoking_DNAm_. Participants were asked how many cigarettes they have smoked in their lifetime, with responses of ‘<5’, ‘5-19’, ’20-49’, ’50-99’ and ‘100+’.

In Generation R, self-reported smoking was obtained via computerized questionnaire during their visit at age 14. Participants indicated whether they had ever smoked (“Have you ever smoked, which could be as little as one cigarette or more than a few puffs?”) and, if applicable, reported the age at first cigarette use (in years and months). Age at first smoking was reverse-coded such that participants who had never smoked were assigned a value of 0, those who first smoked at the oldest reported age (15.0 years) were assigned a value of 1, and each one-year decrease in age at initiation corresponded to an increment of 1 in the score.

In MCS, self-reported smoking was measured at the age 17 sweep. Participants were asked about their smoking habits, with responses of ‘I have never smoked cigarettes’, ‘I have only ever tried smoking cigarettes once’, ‘I used to smoke sometimes but I never smoke a cigarette now’, ‘I sometimes smoke cigarettes now but I don’t smoke as many as once a week’, ‘I usually smoke between one and six cigarettes a week’ and ‘I usually smoke more than six cigarettes a week’.

*Cell Type Proportions*

Estimated counts of cell type (B cells, CD4 T cells, CD8 T cells, granulocytes, monocytes, and natural killer cells) were derived using the Houseman method^17^ with the GSE35069 blood reference panel^18^. In ALSPAC this was done for DNA methylation measured at age 17 and in Generation R this was done for DNA methylation measured at age 18.

*Sex*

In ALSPAC and Generation R, child sex was obtained from midwife and hospital registries.

In MCS, child sex was measured at the age 17 sweep.

*SES Risk Factors*

In ALSPAC, SES risk factor scores – which comprise of mother’s education, experience of reduced income, losing a job, becoming homeless, and financial difficulties – are measured at age 12. Education is coded in order of highest to lowest SES risk: ‘CSE’, ‘Vocational’, ‘O level’, ‘A level’, and ‘Degree’. Experiences of reduced income, homelessness and financial difficulties are also coded in order of highest to lowest SES risk: ‘Affected a lot’, ‘Moderately affected’, ‘Mildly affected’, ‘No effect at all’, and ‘Did not happen’.

In Generation R, SES Risk Factors were defined from maternal education reported by the mother during pregnancy. Educational level ranged from 0 (no education finished) to 5 (university degree obtained) and was reverse coded to indicate SES risk.

In MCS, SES Risk Factors were defined through Organisation for Economic Co-operation and Development (OECD)-equivalised weekly family income at the age 11 sweep. Total net household income was equivalised to account for household size and composition using the modified OECD equivalence scale, assigning a weight of 1.0 to the first adult, 0.5 to additional adults, and 0.3 to each child.

*Batch Effects*

Batch at the DNAm measurement (ALSPAC: 17 years, Generation R: 18 years) were controlled for, as technical factors including batch can lead to variation in methylation levels in blood samples ^19^. In ALSPAC, batch was derived using surrogate variable analysis, estimating 10 surrogate variables which represent unknown factors associated with technical batch ^20^. In Generation R, batch was measured using sample plate number.

*Age at MRI scan*

In ALSPAC, MRI scans were collected over an age range from 18-24 years. In Generation R, MRI scans were collected around the age of 14.

*MRI study*

In ALSPAC, MRI data was collected from three neuroimaging studies ^21^, and so MRI study was controlled for in order to account for variability in study effects.

Supplementary Section 1.4

In ALSPAC, blood samples from young people at age 17 years were selected for analysis as part of the Accessible Resource for Integrative Epigenomic Studies (ARIES, <http://www.ariesepigenomics.org.uk/>;^22^). Bisulfite conversion was performed with the EZ-96 DNA methylation kit (shallow; Zymo Research Corporation, Irvine, USA). DNA methylation levels were then measured on two arrays: the Illumina Infinium HumanMethylation450 and EPIC BeadChip array (Illumina Inc., San Diego, USA). Preprocessing in ALSPAC was performed with the meffil package, using functional normalization. Quality control checks included mismatched genotypes, mismatched sex, incorrect relatedness, low concordance with other time points, extreme dye bias, and poor probe detection. MRIs were conducted using a General Electric 3T HDx scanner. T1-weighted images were processed using the automated FreeSurfer brain imaging software package (Version 6.0.0). Reconstructed images were subjected to quality control measures following the ENIGMA consortium structural image processing protocol (<https://enigma.ini.usc.edu/protocols/imaging-protocols/>; ^21^).

In Generation R, DNA was extracted from peripheral blood samples taken at age 18 years. Five-hundred nanograms of DNA per sample underwent bisulfite conversion using the EZ-96 DNA Methylation kit (Shallow) (Zymo Research Corporation, Irvine, USA) and further processed with the Illumina Infinium MethylationEPIC v2.0 BeadChip. Preprocessing in Generation R was performed with the meffil package, using functional normalization. Magnetic resonance imaging (MRI) data were collected at 14 years of age on a 3T GE MR750W scanner using an eight-channel receive-only head coil (General Electric, Milwaukee, WI). T1-weighted structural images were acquired using a coronal inversion recovery Fast Spoiled Gradient Recalled (IR-FSPGR) sequence, with GE BRAVO option (voxel size = 1.0 mm3 isotropic, TR = 8.77 ms, TE = 3.4 ms, TI = 600 ms, flip angle = 10°, matrix size = 220 × 220, field of view = 220 × 220 mm, slice thickness = 1 mm, number of slices = 230, ARC acceleration factor = 2) ^23^. FreeSurfer v6.0 analysis suite was used for a processing pipeline (https://surfer.nmr.mgh.harvard.edu/), including the removal of non-brain tissue, correction for B1 field inhomogeneities, and whole-brain tissue segmentation ^24^. FSL was used for reorientation and registration to MNI space ^25^. Reconstructed images underwent a rigorous quality control protocol as previously described ^26^. Trained raters visually inspected each scan for inaccuracies in white and pial surface reconstruction, artifacts, and segmentation errors. Images were rated using a five-point Likert scale (unusable, poor, sufficient, good, excellent). Scans rated “sufficient” or better were included in the analyses. Common reasons for exclusion included motion-related artifacts, skull-stripping failures, or pial surface inaccuracies. This quality control protocol followed established procedures and has been implemented in large-scale consortia projects ^27^.

Supplementary Section 1.5

Missing data imputation

In ALSPAC, Generation R and the Millennium Cohort Study, the MICE R package ^28^ was used to impute missing values in the built environment, covariates including SES risk factors, smoking_DNAm_, and age at measurement and cognitive functioning outcomes. The Random Forest method was used for imputation of these variables, and participants were excluded from each analysis if they were missing the relevant biological outcome variable (i.e., DNA methylation or structural MRI) or if they had greater than 50% missingness in the built environment variable. Thirty imputed datasets were generated, with 60 iterations per imputation. Random-forest imputation was implemented using the ranger package with 10 trees for variables assigned the random-forest imputation method. A prespecified predictor matrix was used to determine the variables included as predictors within each imputation model.

In ALSPAC and Generation R, a more conservative threshold of no more than 30% missingness across CpGs (for each clock) for individuals was applied for DNA methylation data, due to the more complex and high-dimensional nature of DNA methylation data ^29^. EpiAge_Altum_ requires 20,318 CpGs, EpiAge_Horvath_ requires 353 and EpiAge_PACE_ requires 173. CpGs were ordered with respect to their chromosomal position, and then imputed by using the k-nearest neighbours (KNN) method, as nearby CpGs often correlate in their methylation levels ^30^. In addition, values more than three times the IQR below the first quartile or above the third quartile were considered outliers. These outlier values were winsorized (replaced with the next lowest/highest valid value - lower or upper bound). This procedure reduces the influence of extreme values while retaining the full dataset for downstream analysis. Completely missing CpG sites were imputed according to epigenetic age package defaults.

In ALSPAC and Generation R, structural MRI data was imputed using ‘MissForest’, a non-parametric method using a Random Forest model ^31^. This was selected as it outperforms KNN methods in contexts whereby data is often on different scales, such as in imaging data ^31^.

Auxiliary variables (i.e., variables selected to inform imputation but not used in the analysis) included:

- Mothers’ travel behaviour at 2-year follow-up (and age at measurement) (ALSPAC only)
- Mothers’ physical activity (at study enrolment) (ALSPAC only)
- Child travel behaviour at age 13 (and age at measurement) (ALSPAC and MCS)
- Child physical health (at age 13) (ALSPAC only)
- Mothers’ depressive symptoms at study enrolment, 4-year follow-up and 18-year follow-up (and age at measurements) (ALSPAC, Generation R – only at study enrolment, and MCS – only at study enrolment)
- Mothers’ familial depressive history (ALSPAC only)
- Mothers’ length of time at address (ALSPAC and MCS)

Auxiliary variables were variables used in previous analyses, have been identified as possible mediators or are closely associated with variables in this study in order to reduce bias ^32^.

**SM Table S4.**

*Built Environment Factor Loadings compared between ALSPAC and Generation R across the three analytic models.*

| Sample | Indicator | Built Environment Factor Loadings | |
| --- | --- | --- | --- |
|  |  | ALSPAC | Generation R |
| Brain Age Models | Building Density | 0.72 | 0.61 |
|  | Connectivity Density | 0.84 | 0.72 |
|  | Facility Richness | 0.71 | 0.83 |
|  | Green Space | -0.79 | -0.61 |
|  | Population Density | 0.54 | 0.43 |
|  | Walkability | 0.77 | 0.83 |
| Epigenetic Age Models | Building Density | 0.76 | 0.61 |
|  | Connectivity Density | 0.82 | 0.72 |
|  | Facility Richness | 0.71 | 0.83 |
|  | Green Space | -0.79 | -0.61 |
|  | Population Density | 0.57 | 0.43 |
|  | Walkability | 0.76 | 0.83 |
| Cognition Models | Building Density | 0.72 | 0.61 |
|  | Connectivity Density | 0.83 | 0.72 |
|  | Facility Richness | 0.73 | 0.82 |
|  | Green Space | -0.77 | -0.61 |
|  | Population Density | 0.52 | 0.43 |
|  | Walkability | 0.78 | 0.83 |
